# Association of public care in childhood with social, criminal, cognitive, and health outcomes in middle-age: six decades of follow-up of members of the 1958 Birth Cohort Study

**DOI:** 10.1101/2020.03.03.20030718

**Authors:** Tiffany H. Xie, Carlos de Mestral, G. David Batty

## Abstract

**Objectives:** To examine if there is an association between childhood public care and adverse life outcomes in middle-age.

**Methods:** We used data from the United Kingdom 1958 birth cohort study of 18,558 babies. Parents of study members were surveyed at age 7, 11, and 16 years when experience of public care of their offspring was ascertained. An array of outcomes were self-reported by cohort members at age 42 years, and a cognitive test battery was administered at age 50.

**Results:** 420 (3.8%) of 11,160 individuals in the analytical sample experienced childhood public care prior to age 16. Net of confounding factors, public care was linked to half of the twenty-eight non-mutually exclusive endpoints captured in middle-age with the most consistent effects apparent for psychosocial characteristics: 6/7 sociodemographic, 2/2 anti-social, 3/3 psychological, 1/3 health behaviours, 2/8 somatic health, and 0/5 cognitive.

**Conclusions:** The present study suggests that known associations between childhood care and outcomes in adolescence and early adulthood are also seen in middle-age.

**Policy implications:** Practitioners in health and social services should perhaps more closely monitor care graduates.

## Introduction

Prospective and retrospective cohort studies conducted over several decades have repeatedly demonstrated associations of childhood socioeconomic disadvantage with adult mortality and cardiovascular disease.^1,2^ Such early life adversity has recently been more broadly defined to not only comprise economic hardship but also physical and emotional abuse, nutritional privation, sibling loss, parental separation, and out-of-home care, amongst other characteristics.^3^ Children removed from their home into state care – also referred to as being looked after or public care – are typically placed with a foster family and usually separated from their biological families in order to protect their welfare following parental abuse or neglect, or, less commonly, as a result of their own anti-social behaviour. In recent years there has been a secular rise in the number of children placed in care in both the US^4^ and the UK,^5^ with current estimates suggesting up to 4% of the UK population may have experience of pre-adult care.

Although public care is predicated upon providing a more nurturing and protective environment for children than available via their family of origin, there is emerging evidence that, in fact, exposure to childhood care is linked to poor outcomes. As such, relative to their unexposed peers, children with a history of public care followed into late adolescence and early adulthood are more likely to experience a greater prevalence of teenage pregnancy,^6^ school exclusion,^7^ mental health problems, particularly depression,^8,9^ and substance abuse,^7,10^ and be implicated in criminal activity.^9,11^ These unfavourable outcomes are apparent alongside substantial socioeconomic disadvantage as characterised by poverty,^12^ an absence of health insurance in societies without universal coverage,^13^ low educational attainment, and insecure housing tenure.^11^ Notably, many of these effects appear to hold after taking into account early life confounding factors including illness and disability, suggesting that the impact of care may be independent of other adverse psychosocial characteristics that tend to prevail at the time of care placement.

While it might be anticipated that this array of shorter-term negative life outcomes will position looked after children on a negative life trajectory into middle-age, studies exploring the longer term impact of early life care are very sparse. Most investigators have used a cross-sectional design in which participants were asked to recall care as much as six decades after the experience,^9,11,12,14^ so raising concerns regarding recall bias. In the only prospective cohort study of which we are aware, economic hardship and mental health problems were associated with prior care exposure in a large cohort of Swedish children resurveyed at age 55 years,^15^ but the impact of care on a broader array of outcomes is untested.

We therefore addressed this gap in knowledge using data from a six decade-long follow-up of members of a large British birth cohort study in whom care data was prospectively gathered across childhood. This study also holds information on harmful health behaviours, such as heavy drinking and smoking, psychological problems, anti-social conduct, and, uniquely in the context research on the consequences of care, cognitive function. These data allow us to test the hypothesis that gradients between childhood public care and outcomes apparent in early adulthood will largely extend into middle-age.

## Methods

We used data from the National Childhood Development Study, also known as the 1958 Birth Cohort Study.^16,17^ An ongoing, closed, prospective cohort study – 11 surveys have been conducted to date – investigators have followed 18,558 births occurring in the United Kingdom during a single week in the late 1950s. In the present analyses we used data from birth to 16 years of age to capture public care and potential covariates, while outcomes were assessed at surveys which took place at ages 42 and, for cognitive function only, 50 years. In childhood, participant consent was obtained from parents or caregivers, and, later, the study members themselves. The study was originally approved by the National Health Service Research Ethics Committee and latterly the Multicentre Research Ethics Committee. The present report conforms to STROBE (STrengthening the Reporting of OBservational studies in Epidemiology) guidelines.^18^

### Assessment of out-of-home care and covariates

We ascertained childhood history of public care from prospective surveys conducted when participants were aged 7, 11, and 16 years. Parents were asked: “Has the child been under the care of the local authority, now or in the past?” Study members were regarded has having experienced care in childhood if a positive response was provided to enquiries at any of the three surveys. We derived three indicators of childhood socioeconomic status as recorded at birth: paternal occupational social class as based on the Registrar General’s social classification of occupations (6 levels),^19^ maternal education level denoted by age at leaving full-time educations (in the era of data collection, school was mandatory until age 15 years), and number of persons in the household. Indicators of childhood health, recorded at age 7, included hospital visits for illnesses or medical tests, any disability (including birth defects, visual/hearing/speech handicaps, epilepsy, and asthma), and internalizing and externalizing symptoms, as quantified using the Bristol Social Adjustment Guides.^20,21^ Summary scores were created for the internalizing symptoms of unforthcomingness, withdrawal, depression, and miscellaneous symptoms, and for the externalizing symptoms of anxiety for acceptance by adult or peers, hostility towards adults, restlessness, and inconsequential behaviour. Remaining covariates were maternal age at birth of study member and maternal marital status at birth (married, separated, divorced, widowed, stable union, and unmarried).

### Assessment of adult outcomes

All adult characteristics were self-reported. At age 42, we assessed occupational social class (as above), unemployment, receipt of any social benefits by the participant or their partner, educational attainment, cohabitation, marital status, and history of homelessness (since age 33 years). Indicators of anti-social behaviour were use of illegal drugs in the past year and having received a criminal conviction (since age 33 years). We quantified psychological distress using the Malaise Inventory.^22^ Validated in the general population, including the present sample,^23^ this is a 24-item scale in which a score of ≥7 has been used to indicate morbidity. In addition to the reporting of symptoms, diagnosis was captured using enquiries about being under the care of a medical specialist for depression or being hospitalised in the past year for its treatment. Alcohol problems were assessed using the CAGE (‘Cutting down, Annoyance by Criticism, Guilty feeling, and Eye-openers’) Questionnaire. This is also validated screening instrument where a score of ≥2 indicates possible alcohol dependence or abuse,^24,25^ a threshold that has been shown to have predictive capacity for mortality.^26^

Current smoking was based on standard enquiries. For alcohol intake, study members were asked how many drinks they had consumed in the past 7 days, including beer, wine, spirits, sherry, ‘alcopops’, and any other alcoholic drinks. Heavy drinking was denoted by exceeding specific thresholds for women (>35 drinks/week) and men (>50). Participants were asked how often they take part in any physical activity with low levels being defined by exercising fewer than 3 times a week.^27^ Obesity was denoted by a body mass index of ≥30 kg/m^2^ as derived from self-reported height and weight.^28^ Study members reported a physician diagnosis of diabetes and high blood pressure. Other health indices included physical disability – denoted by permanent illness or injury for six or more months – accidents (since age 33 years), hospital admissions, occurrence of any cancer, and general health. General health was rated on a standard four-point scale (excellent, good, fair, or poor). Accidents were defined as events such as road- or sports-related injury, sexual assault, or muggings that required medical attention.

Lastly, cognitive function at age 50 was measured with a battery of memory and executive functioning tests which have been widely used in other population-based surveys.^29^ Memory was assessed by a test which involved memorising words with immediate and delayed recall. Executive functioning was measured by letter cancellation and animal naming tasks.

### Statistical analysis

We used odds ratios with accompanying confidence intervals, computed using logistic regression, to summarise the relationship between childhood care and outcomes at age 42 years, while linear regression models produced beta coefficients, also with 95% confidence intervals, to quantify the relationship between care and indicators of cognitive function at 50 years. For all endpoints, effect estimates were first adjusted for sex and then for a series of covariates which characterised the early life socio-demographic and health circumstances of the study member; as a birth cohort study, controlling for age was not required. All data analyses were conducted using Stata SE (Version 15.1, College Station, TX: StataCorp).

## Results

In **Figure 1** we show the flow of study members into the analytical sample. In total, care history data across childhood were available for 16,583 study members; of these, 11,160 participated in the survey at age 42 and 9578 in the resurvey at age 50 years. Those study members who did not take part in the age 42 sweep were somewhat more likely to have experienced care in pre-adulthood (5.6%) than participants (3.8%). The same observations were made at age 50.

**Figure 1.**
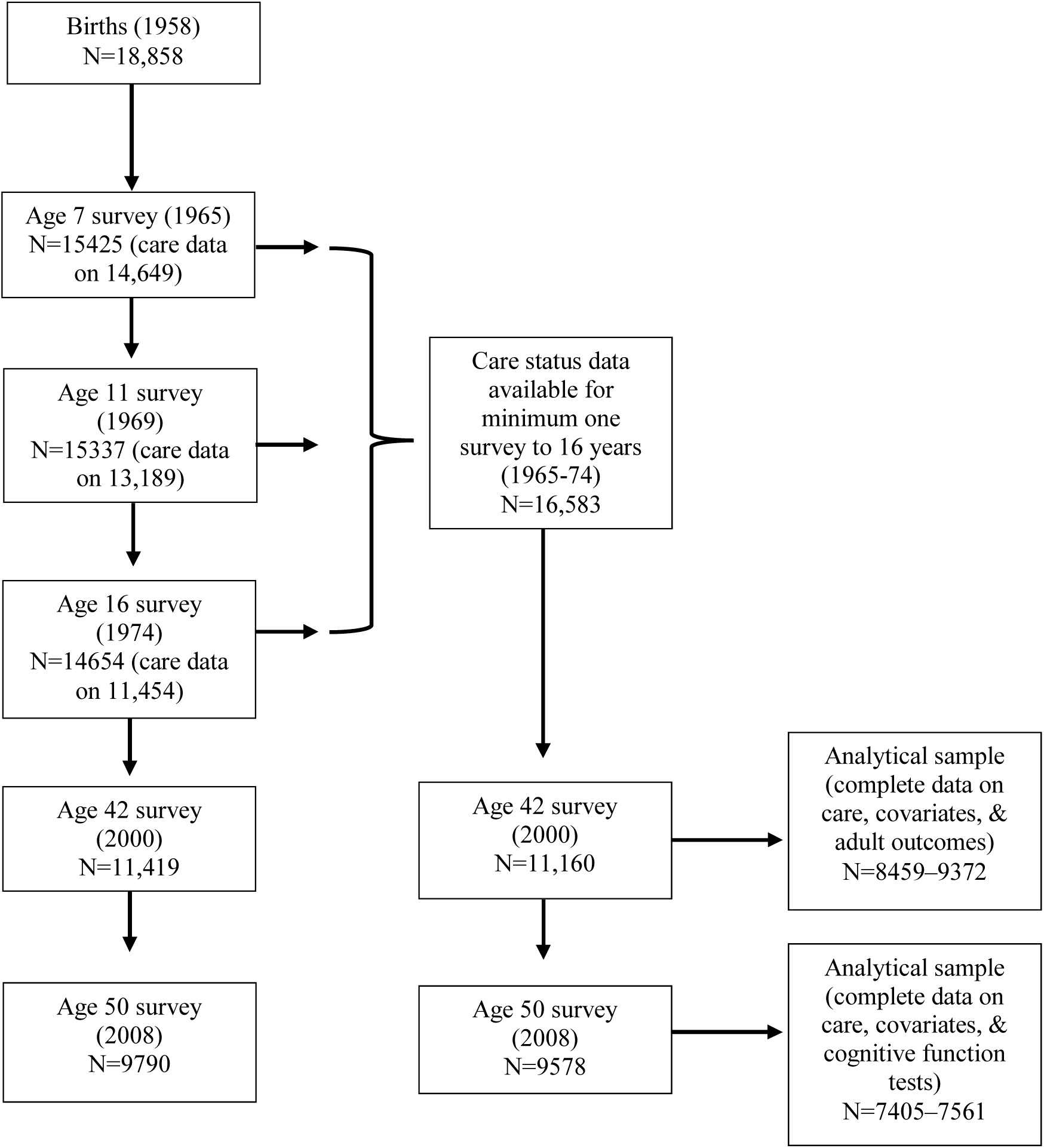
Flow of study members through phases of data collection into the present analytical sample, 1958 Birth Cohort Study.

In **Table 1**, we present study member early life characteristics according to history of childhood care contingent upon participation in the survey at age 42 years. In this analytical sample, 420 (3.8%) had a history of care before age 16. In general, children who had a history of being in public care had markedly less favourable characteristics relative to their unexposed peers. Thus, exposed children were more likely to come from socioeconomically disadvantaged backgrounds as evidenced by a greater prevalence of overcrowding, manual paternal social class, and lower maternal education. Similarly, such children had a higher health burden based on hospital visits, disability, and internalizing and externalizing symptoms, the latter differences being particularly marked. Given the clear patterning of early life characteristics according to public care exposure, we incorporated these factors as confounders in the following multivariable regression models in which we attempted to ascertain the independent relation, if any, between childhood care and adult outcomes.

**Table 1.**
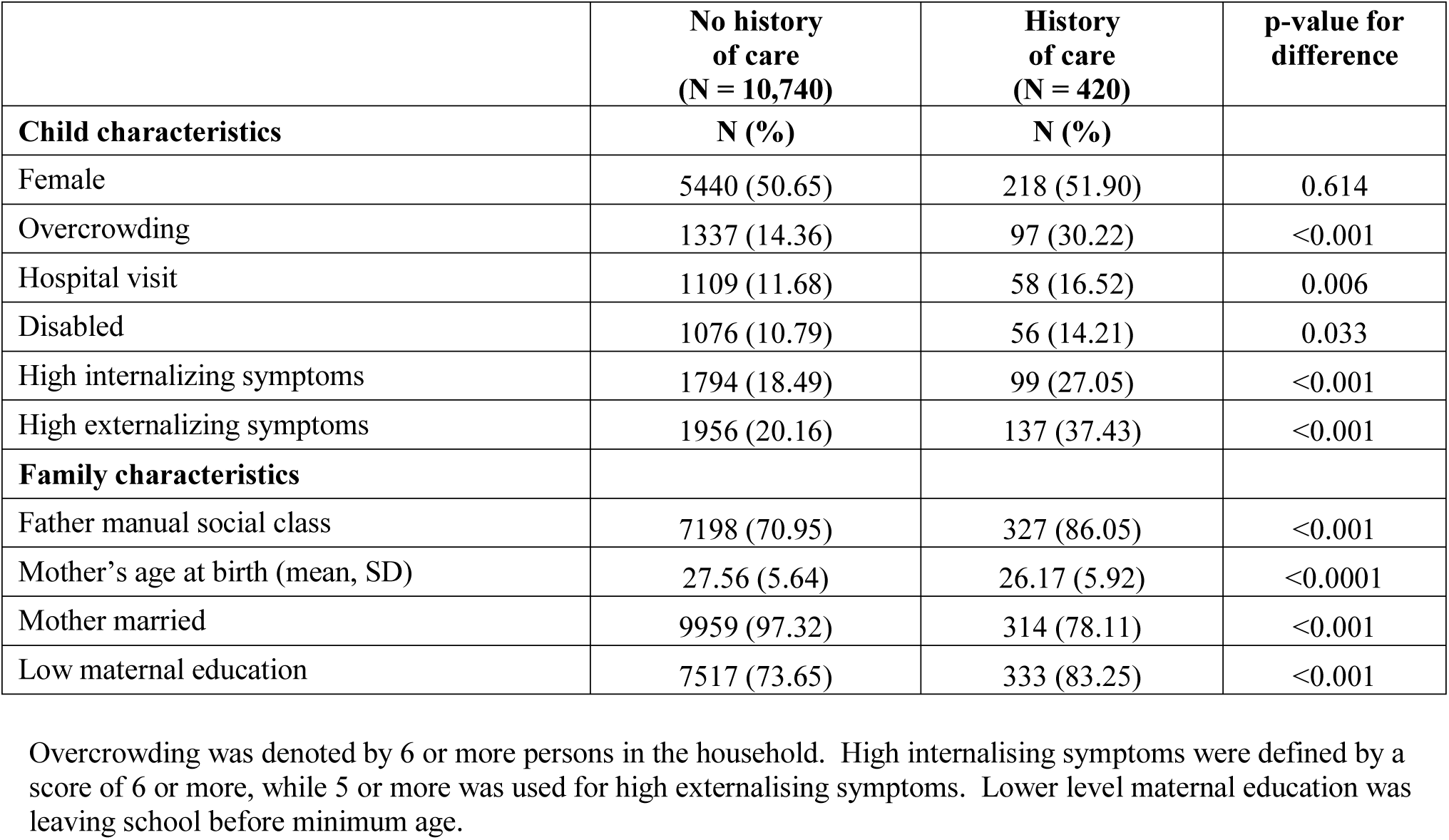
Early life characteristics of study members according to history of childhood care, 1958 Birth Cohort Study.

|  | No history<br>of care<br>(N = 10,740) | History<br>of care<br>(N = 420) | p-value for<br>difference |
| --- | --- | --- | --- |
| **Child characteristics** | **N (%)** | **N (%)** |  |
| Female | 5440 (50.65) | 218 (51.90) | 0.614 |
| Overcrowding | 1337 (14.36) | 97 (30.22) | <0.001 |
| Hospital visit | 1109 (11.68) | 58 (16.52) | 0.006 |
| Disabled | 1076 (10.79) | 56 (14.21) | 0.033 |
| High internalizing symptoms | 1794 (18.49) | 99 (27.05) | <0.001 |
| High externalizing symptoms | 1956 (20.16) | 137 (37.43) | <0.001 |
| **Family characteristics** |  |  |  |
| Father manual social class | 7198 (70.95) | 327 (86.05) | <0.001 |
| Mother's age at birth (mean, SD) | 27.56 (5.64) | 26.17 (5.92) | <0.0001 |
| Mother married | 9959 (97.32) | 314 (78.11) | <0.001 |
| Low maternal education | 7517 (73.65) | 333 (83.25) | <0.001 |

Overcrowding was denoted by 6 or more persons in the household. High internalising symptoms were defined by a score of 6 or more, while 5 or more was used for high externalising symptoms. Lower level maternal education was leaving school before minimum age.

In **Table 2** we depict the association of experience of public care with sociodemographic, anti-social, and psychological outcomes at age 42 years. Following adjustment for sex, individuals with a history of childhood care were more than twice as likely to be in a manual social class, be unemployed, be in receipt of social benefits, have only a basic education, and have experienced homelessness in adulthood compared to unexposed individuals. Children from a care background were also markedly more likely to live alone several decades later, though there was no apparent link with marital status. Separate control for potential confounding factors from early life, particularly poverty rather than indices of health, led to partial attenuation for some but not all associations of childhood care with adult outcomes. Statistical significance at conventional levels was typically retained, however, as evident by the absence of unity in the accompanying confidence intervals.

**Table 2.** Association of care in childhood with sociodemographic, anti-social, and psychological outcomes at age 42 years, 1958 Birth Cohort Study.

|  | Sex-adjusted |  | Sociodemographic-adjusted |  | Health-adjusted |  |
| --- | --- | --- | --- | --- | --- | --- |
|  | OR (95% CI) | N | OR (95% CI) | N | OR (95% CI) | N |
| **Sociodemographic** |  |  |  |  |  |  |
| Manual social class | 2.14 (1.75, 2.62) | 11139 | 1.60 (1.25, 2.05) | 9251 | 1.89 (1.51, 2.38) | 9565 |
| Unemployed | 2.49 (1.57, 3.93) | 11126 | 1.62 (0.86, 3.07) | 9249 | 1.78 (1.01, 3.11) | 9565 |
| Social benefits | 2.65 (2.08, 3.37) | 11096 | 2.41 (1.80, 3.23) | 9222 | 2.39 (1.82, 3.14) | 9539 |
| Secondary education or less | 2.11 (1.64, 2.73) | 11786 | 1.43 (1.03, 1.99) | 8614 | 1.86 (1.36, 2.53) | 8884 |
| Living alone | 1.66 (1.23, 2.25) | 11125 | 1.63 (1.14, 2.35) | 9245 | 1.58 (1.13, 2.20) | 9557 |
| Never married | 1.16 (0.83, 1.61) | 11102 | 1.01 (0.67, 1.52) | 9228 | 1.02 (0.70, 1.48) | 9539 |
| Homelessness | 2.18 (1.60, 2.99) | 10569 | 2.23 (1.54, 3.24) | 8800 | 2.00 (1.40, 2.85) | 9089 |
| **Anti-social** |  |  |  |  |  |  |
| Illegal drugs | 2.32 (1.51, 3.57) | 11009 | 2.14 (1.26, 3.64) | 9158 | 2.27 (1.41, 3.64) | 9471 |
| Criminal conviction | 2.15 (1.59, 2.91) | 11020 | 1.64 (1.13, 2.38) | 9167 | 1.71 (1.22, 2.40) | 9481 |
| **Psychological** |  |  |  |  |  |  |
| Psychological distress | 1.85 (1.48, 2.32) | 11008 | 1.64 (1.25, 2.16) | 9157 | 1.85 (1.45, 2.37) | 9470 |
| Specialist care for depression | 2.49 (1.74, 3.55) | 11036 | 1.82 (1.13, 2.93) | 9180 | 2.24 (1.49, 3.36) | 9493 |
| Alcohol problems | 1.47 (1.13, 1.91) | 10961 | 1.10 (0.78, 1.53) | 9130 | 1.37 (1.02, 1.85) | 9438 |

OR, odds ratio. Sociodemographic-adjusted odds ratios are adjusted for sex plus parental social class, mother's age at birth, mother's marital status, mother's education, and high number of people in household. Health-adjusted odds ratios are adjusted for sex plus childhood hospitalizations, childhood disability, and internalizing and externalizing symptoms.

Compared to participants who had not been in public care, those with such a history were more likely to engage in delinquency in middle-age, as evidenced by a doubling of risk of illicit drug use and a criminal conviction. While taking into account early social circumstances diminished the magnitude of effects for criminality, this was not the case for drug use.

Participants with a history of care were more than twice as likely as those without to have had a diagnosis of adult depression, as denoted by being under the care of a specialist and/or being hospitalised. Notably, while controlling for socioeconomic position in childhood led to some attenuation of these odds ratio but this was not the case after taking into account childhood distress, amongst other health characteristics. The care–alcohol problems relationship was more marked than for drinking behaviour (**Table 3**). Indeed, with the exception of cigarette smoking, where a doubling in prevalence was apparent in adults who, as children, had been looked after, the health behaviours of alcohol consumption and physical exertion were not strongly linked to care status.

**Table 3.** Association of care in childhood with health behaviours and somatic health outcomes at age 42 years, 1958 Birth Cohort Study.

|  | Sex-adjusted |  | Sociodemographic-adjusted |  | Health-adjusted |  |
| --- | --- | --- | --- | --- | --- | --- |
|  | OR (95% CI) | N | OR (95% CI) | N | OR (95% CI) | N |
| **Health behaviours** |  |  |  |  |  |  |
| Daily smoker | 2.49 (2.04, 3.03) | 11119 | 1.77 (1.39, 2.25) | 9244 | 2.36 (1.89, 2.95) | 9560 |
| Heavy alcohol use | 1.47 (0.89, 2.43) | 11147 | 1.19 (0.63, 2.25) | 9258 | 1.16 (0.63, 2.15) | 9571 |
| Physically inactive | 0.92 (0.75, 1.11) | 11116 | 0.95 (0.75, 1.20) | 9242 | 0.89 (0.72, 1.12) | 9558 |
| **Somatic health** |  |  |  |  |  |  |
| Obese | 0.94 (0.71, 1.25) | 11160 | 0.77 (0.54, 1.08) | 9266 | 0.92 (0.67, 1.25) | 9581 |
| Diabetes | 0.69 (0.28, 1.68) | 11120 | 0.44 (0.14, 1.42) | 9244 | 0.62 (0.23, 1.70) | 9560 |
| High blood pressure | 1.12 (0.84, 1.51) | 11116 | 1.28 (0.91, 1.80) | 9240 | 1.16 (0.84, 1.61) | 9556 |
| Disabled | 3.55 (2.72, 4.62) | 11160 | 2.98 (2.16, 4.11) | 9266 | 3.06 (2.25, 4.16) | 9581 |
| Hospital admission | 1.10 (0.90, 1.36) | 11115 | 1.18 (0.92, 1.50) | 9240 | 1.20 (0.95, 1.50) | 9557 |
| Accident | 1.33 (1.09, 1.63) | 11132 | 1.27 (0.99, 1.62) | 9252 | 1.14 (0.91, 1.43) | 9570 |
| Any cancer type | 1.71 (1.02, 2.86) | 11120 | 1.57 (0.83, 2.97) | 9244 | 1.86 (1.06, 3.24) | 9560 |
| Poor general health | 1.68 (1.34, 2.10) | 11119 | 1.43 (1.09, 1.88) | 9243 | 1.66 (1.29, 2.12) | 9559 |

OR, odds ratio. Sociodemographic-adjusted odds ratios are adjusted for sex plus parental social class, mother's age at birth, mother's marital status, mother's education, and high number of people in household. Health-adjusted odds ratios are adjusted for sex plus childhood hospitalizations, childhood disability, and internalizing and externalizing symptoms.

Individuals with a history of care were almost three times as likely to experience physical disability in adulthood compared to individuals without such a background. These individuals were also more likely to report poor general health and cancer from all sites combined. There were no clear link between pre-adult care and later obesity and, perhaps as a result, relationships with adult diabetes and hypertension were not apparent.

In **Figure 2** we show the results from the multivariable models in which we adjust the association between care and adult outcomes for sex, sociodemographic and health covariates from childhood. Most of the socioeconomic outcomes were robust to such statistical treatment. Other endpoints which were also related to public care based on these more complex models included adult mental health, illicit drug use, smoking, disability and poor self-rated health.

**Figure 2.**
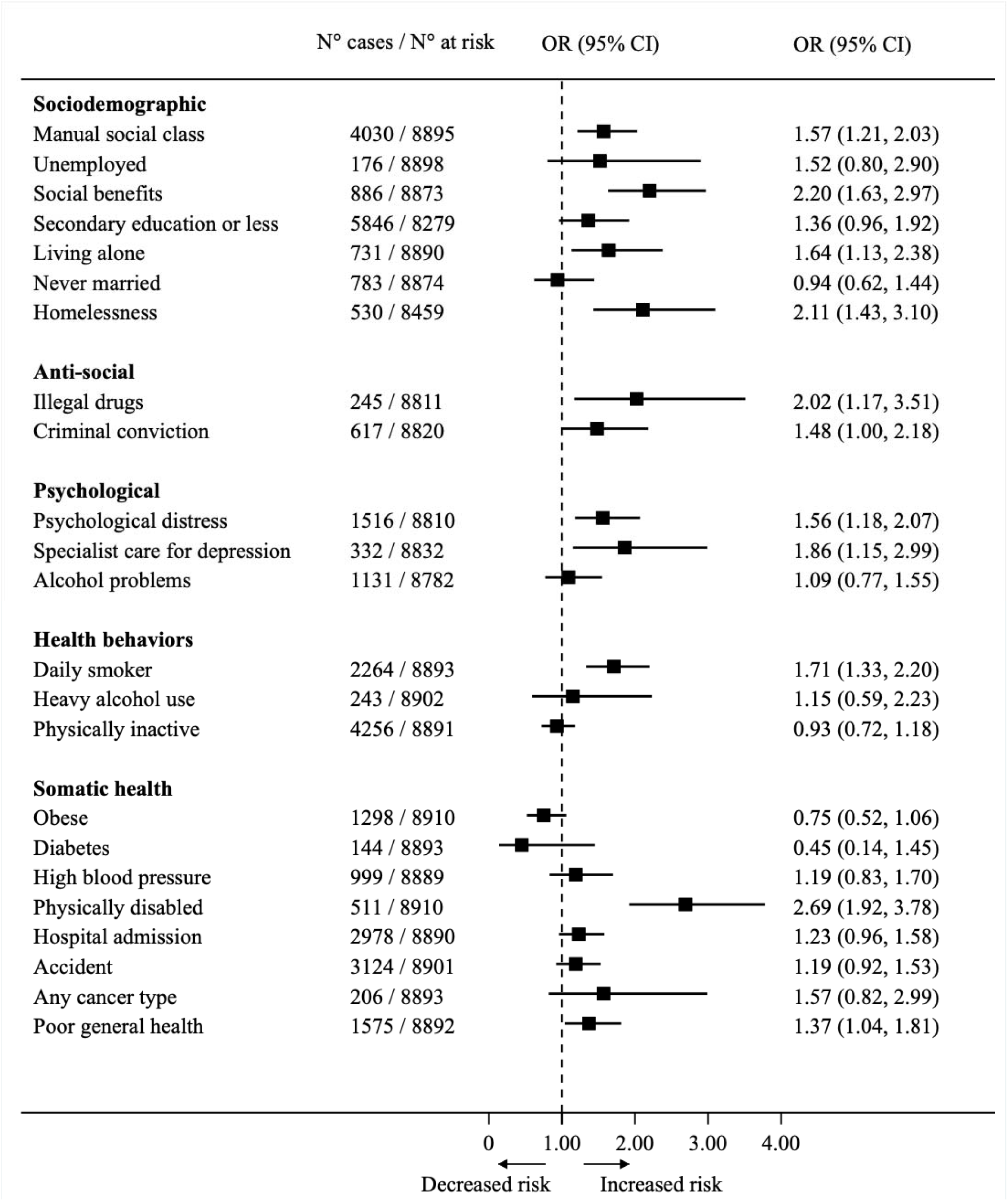
Multivariable adjusted association between care in childhood and outcomes at age 42 years, 1958 Birth Cohort Study. Multiply-adjusted odds ratios (OR) and 95% confidence intervals (CI) are adjusted for the following early life factors: sex, parental social class, mother’s age at birth, mother’s marital status, mother’s education, high number of people in household, childhood hospitalizations, childhood disability, and internalizing and externalizing symptoms.

We also examined if being looked after in early life was related to cognitive function at age 50 years (**Table 4**). The mean difference in performance on 5 tests according to care status was modest. With the exception of letter cancellation speed, an indicator of sustained concentration, these differences were apparent after sex-adjustment but were largely lost after control for multiple potential confounding factors. Lastly, when we repeated all our analyses based on individuals without missing data, our conclusions were largely unchanged for both non-cognitive (**Supplemental Table 1**) and cognitive outcomes (**Supplemental Table 2**).

**Table 4.** Association of care in childhood with cognitive function at age 50 y, 1958 Birth Cohort Study.

|  | No history of<br>care | History of<br>Care | Sex-adjusted |  | Sociodemographic-adjusted |  | Health-adjusted |  | Multiply-adjusted |  |
| --- | --- | --- | --- | --- | --- | --- | --- | --- | --- | --- |
|  | Mean (SD) | Mean (SD) | B (95% CI) | N | B (95% CI) | N | B (95% CI) | N | B (95% CI) | N |
| Letter Cancellation Accuracy | 21.65 (5.87) | 20.90 (6.50) | -0.81 (-1.45, -0.16) | 9243 | -0.89 (-1.67, -0.10) | 7705 | -0.75 (-1.48, -0.02) | 7953 | -0.83 (-1.64, -0.03) | 7405 |
| Letter Cancellation Speed | 334.39 (88.41) | 330.01 (98.33) | -5.1 (-14.79, 4.59) | 9243 | -8.25 (-20.08, 3.58) | 7705 | -6.13 (-17.18, 4.92) | 7953 | -9.28 (-21.49, 2.93) | 7405 |
| Animal Naming | 22.29 (6.29) | 21.41 (6.51) | -0.88 (-1.57, -0.20) | 9444 | 0.06 (-0.76, 0.87) | 7870 | -0.53 (-1.29, 0.23) | 8120 | 0.23 (-0.60, 1.07) | 7561 |
| Word Recall Immediate | 6.56 (1.49) | 6.23 (1.47) | -0.33 (-0.49, -0.17) | 9444 | -0.17 (-0.36, 0.03) | 7870 | -0.20 (-0.38, -0.02) | 8120 | -0.12 (-0.32, 0.08) | 7561 |
| Word Recall Delayed | 5.43 (1.84) | 5.01 (1.88) | -0.43 (-0.63, -0.24) | 9388 | -0.22 (-0.46, 0.02) | 7823 | -0.29 (-0.51, -0.07) | 8071 | -0.15 (-0.39, 0.10) | 7514 |

B, beta coefficient. Sociodemographic-adjusted odds ratios are adjusted for sex plus parental social class, mother's age at birth, mother's marital status, mother's education, and high number of people in household. Health-adjusted odds ratios are adjusted for sex plus childhood hospitalizations, childhood disability, and internalizing and externalizing symptoms. Multiply-adjusted odds ratios are adjusted for all these covariates.

## Discussion

The main findings of this study were that a history of public care during childhood was associated with a broad range of negative outcomes in adulthood, most consistently socioeconomic factors and anti-social behaviour. That is, after multivariable adjustment and at conventional levels of statistical significance, public care was linked to half of the thirty endpoints featured in the present analyses, comprising 6/7 sociodemographic, 2/2 anti-social, 3/5 psychological, 1/3 health behaviours, 2/8 somatic health, and 0/5 cognitive.

While these findings may be important given the pernicious nature of several of the outcomes in their own right, they may also offer insights into the observation that, relative to the general population and net of confounding, children who are exposure to care may experience up to three times the rate of premature mortality by middle-age.^30-32^ This observation raises the question of how exposure to public care in childhood is embodied, in turn elevating mortality. In analyses of two cohort studies, we have recently shown little evidence of an association between pre-adult public care and various adult cardiovascular, inflammatory, neuroendocrine, and respiratory risk markers for mortality.^33,34^ While the present results suggest that, instead, the impact of public care on death may be via social circumstances and potentially some indicators of health, our study sample is insufficiently mature to enable us to simultaneously run analyses in which we incorporate data on exposure, potential mediators, and mortality risk. In the only study of which we are aware to have done so, indices of socio-economic position and mental health appear to have some explanatory role in the public care–mortality relationship.^35^ Replication and testing using a wider range of potential mediating characteristics, such as somatic health, health behaviours and criminality as identified herein, is now required.

### Comparison with existing studies

Our results of a wide-ranging association of pre-adult care with outcomes in middle-age broadly accord with evidence from samples drawn from Scandinavia,^8^ US,^12^ and UK^9^ populations followed earlier in the adult life course. Our findings also seem to corroborate those from cross-sectional studies including older-aged groups that relied on distant recall of early life care.^9,11,12,14^

In the present analyses, while childhood care was associated with a higher burden of mental health problems there were less consistent relationships with health behaviours and somatic health outcomes. Thus, our findings support analyses of data from the 1970 British birth cohort which found associations between care history and smoking, but not alcohol problems or physical exertion.^36^ We did not find associations between care history and either high blood pressure or obesity, although these are not universal observations.^13,14^

To our knowledge, ours was the first study to explore the influence of being looked after in childhood on cognition in mid-life. With both care^33^ and cognition^37^ being strongly socially patterned, we anticipated that care would be related to lower cognitive performance but this was not the case – even after basic adjustment, the difference in performance on various tests of mental ability according to care status was marginal.

### Study strengths and limitations

The strengths of our study include the well-characterised study participants, the prospective measurement of public care across childhood as opposed to distant recall in older age, and our use of an extensive range of outcomes which, unusually in the context of the present literature, allowed us to compare effects within a single population. Our study is of course not without several limitations. First, 59% of cohort members with care data took part in the resurvey at age 42 (51% at age 50 years), with, as shown, lower participation rates in individuals with such a history. Provided there was still sufficient variation in exposure and outcomes in the remaining study members, and given that our objective was to explore relationships as opposed to examining the prevalence of a given characteristic, attrition should not have impacted upon our findings.^38,39^ Second, parents may provide a socially desirable response to an enquiry as revealing and sensitive as the public care history of their offspring. While we are unaware of any studies assessing the agreement of parental self-reported care with a gold standard such as administrative data, in preliminary findings from a meta-analysis of studies of childhood care exposure and adult mortality risk in which we compared the predictive value of care data as ascertained from parental report with electronic records across studies, we found that care data collected using each mode were associated with around a doubling of adult mortality rates in those with a history of care relative to those without.^40^ This suggests similar predictive validity for self-reported and administrative data. Outcome data were also self-reported, on this occasion by the study members as opposed to parents, but many of these have been shown to sufficiently valid for the purposes of epidemiological investigation.

Third, vulnerable children and families are hard to reach in a research context, and it is likely that the present study will not capture data on people who experienced the more challenging of upbringings. For that, purposeful sampling of such groups would be required alongside unexposed children to facilitate comparison. Fourth, we were unable to examine potentially important care placement characteristics, including the impact of duration, type, reason, or stability. Fifth, although we attempted to take into account the early life circumstances of children experiencing care, it is not possible to completely separate the impact of the various adversities experienced prior to and during care. Finally, the public care system in the UK has evolved in the several decades since the present study members were children. Some changes, most obviously the establishment of the modern UK protection system in 1973, have seemingly improved the circumstances for this disadvantaged population such that they now receive intervention earlier and experience less placement instability.^41^ While this raises the possibility that the apparent longer-term negative effects may be diminished for children in contemporaneous care, there is in fact evidence that more recent generations of children in foster care still experience subsequent adversity.^13,42^

### Public health implications and future research directions

Taken together with the findings of other observational studies, there is growing evidence of risk in multiple domains across the adult life course for people exposure to childhood care. As with all potential population-based interventions, similarly consistent experimental evidence must now be accumulated in order to inform public policy. Our results and those of other groups are likely to be informative for the planning and implementation of trials, ideally randomised with control groups. Given the logistical and financial implications of such an endeavour, however, quasi experimental studies which utilise changes in child protection legislation as an instrumental variable may also have value. In the meantime, practitioners in health and social care should perhaps more closely monitor care graduates.

### Conclusions

This study adds to a growing body of evidence of the shorter-term negative impact of public care in childhood on a broad range of outcomes to suggest that many effects extend further into the adult life course than previously understood.

## Data Availability

Data are available from the Uk Data Archive.

https://www.data-archive.ac.uk/

## Acknowledgements

GDB is supported by the UK Medical Research Council (MR/P023444/1) and the US National Institute on Aging (1R56AG052519-01; 1R01AG052519-01A1). CdM was supported by the Swiss National Science Foundation when this work was initiated.

## Human participant protection

Participant consent was obtained throughout the study from parents or caregivers, and, later, the study members themselves. The study was originally approved by the National Health Service Research Ethics Committee and latterly the Multicentre Research Ethics Committee.

## Contributions

Tiffany Xie analysed the data, interpreted the results, and jointly drafted the manuscript. Carlos de Mestral built the dataset, interpreted the results, and revised the manuscript. David Batty generated the idea for the study, formulated an analytical plan, interpreted the results, and jointly drafted the manuscript. All authors approved the final version of the manuscript.

## Manuscript statistics

3341 words (excluding abstract 185 words), 42 references, 4 tables (2 supplemental tables), 2 figures

